# One size fits all?: Modeling face-mask fit on population-based faces

**DOI:** 10.1101/2020.10.07.20208744

**Authors:** Tomas Solano, Rajat Mittal, Kourosh Shoele

**Affiliations:** Department of Mechanical Engineering, Florida State State University, Tallahassee, FL 32310, USA; Department of Mechanical Engineering, Johns Hopkins University, Baltimore, MD 21218, USA; School of Medicine, Johns Hopkins University, Baltimore, MD 21205, USA

## Abstract

The use of face masks by the general population during viral outbreaks such as the COVID-19 pandemic, although at times controversial, has been effective in slowing down the spread of the virus. The fit of simple cloth masks on the face as well as the resulting perimeter leakage and face mask efficacy are expected to be highly dependent on the type of mask and facial topology. However, this effect has to date, not been examined and quantified. Here, we study the leakage of a rectangular cloth mask on a large virtual population of subjects with diverse facial features, using computational mechanics modeling. The effect of weight, age, gender, and height on the leakage is studied. The Centers for Disease Control and Prevention (CDC) recommended mask size was used as a basis for comparison and was found *not* to be the most effective design for all subjects. Thin, feminine, and young faces benefit from mask sizes smaller than that recommended by the CDC. The results show that side-edge tuck-in of the masks could lead to a larger localized gap opening in many face categories, and is therefore not recommended for all. The perimeter leakage from the face mask worn by thin/feminine faces is mostly from the leakage area along the bottom edge of the mask and therefore, a tuck-in of the bottom edge of the mask or a mask smaller than the CDC recommended mask size are proposed as a more effective design. The leakage from the top edge of the mask is determined to be largely unaffected by mask size and tuck-in ratio, meaning that other mechanical alterations such as a nose wire strip are necessary to reduce the leakage at this site.

## 1 Introduction

Over 200 scientists, have urged public-health authorities to acknowledge the potential for airborne transmission of the novel SARS-CoV-2 coronavirus [3]. While there is still a lot that is unknown about the transmission of the SARS-CoV-2 virus, it is evident now that like its predecessor, SARS-CoV-1, airborne transmission is a significant mode of transmission [4, 5, 26]. Airborne transmission happens when a susceptible person inhales microscopic bio-aerosols in the air which are generated from a respiratory event such as a cough, sneeze, or even just breathing and talking [4, 6]. Aerosols are a suspension of solid or liquid particles in a gas, made up of droplets smaller than 10*μm*. While larger droplets (≥ 100*μm*) reach the ground within a second, aerosols can linger in the air for hours, increasing the probability of a susceptible person coming in contact with the virus [7, 8].

Experimental studies with human subjects and manikins show that mask usage can limit the droplet and airborne transmission of various infections to and from the wearer [9–17]. However, the effectiveness of the outward protection of masks as well as the role of varying mask fit on the face, remain largely unexplored. Air leakage has been observed around the perimeter of the mask where it does not make a seal with the face, reducing the effectiveness of the mask [7]. Perimeter leakage is caused by loose or improper fitting face masks and can be significantly impacted by facial features [18–21]. While proper fitting of N95 respirators on a face has always been stressed for effective filtering of all contaminants, there is a lack of knowledge on how important the fitting of homemade cloth and simple surgical masks is. Surgical masks are primarily designed for outward protection from droplets, not aerosols, and therefore, the fit is much looser. Homemade masks made from cotton or similar fabrics are likely to be even looser, thus allowing for more leakage. These loser fitting masks are more susceptible to perimeter leakage and, therefore, not as effective against aerosols.

In a study comparing the effectiveness of different face masks, homemade masks were shown to be half as effective as surgical masks and 50 times less effective than an FFP2 mask. These effects were even more pronounced amongst the children subjects, likely due to an inferior fit on their smaller faces [16]. More recently, Verma et al. experimentally explored the effect of different mask types by visualizing the respiratory jets and observed leakage through the perimeter of the mask [22]. The study reported that both the mask material and fit have an important impact on the mask’s effectiveness, with all masks tested showing leakage from the top of the mask due to poor fitting. The studies by Oestenstad et al [19] and Oestenstad and Bartolucci [20] used a fluorescent tracer to identify leak location and shape on subjects wearing half-mask respirators. They tested the effect of gender, race, respirator brand, and facial dimensions and found that facial dimensions were positively correlated with leak location. Tang et al studied the jet generated from coughing and the effect of wearing surgical masks or N95 respirators [23]. They found that a surgical mask effectively blocks the forward momentum of the cough jet, but the loose fit of the mask allows air leakage around the perimeter of the mask primarily through the top and sides. Lei et al. used a headform finite element model to study the leakage locations of an N95 respirator and show that the most leakage occurs along the top perimeter of the mask near the nose [24]. The simulations of face-masks and population-based headform models have the potential to accurately estimate the location and amount of leakage for different facial structures. This can lead to better transmission models for future viral outbreaks and more accurate predictions of the face-masks efficacy on different people.

A strong argument can be made for the importance of accurate mask-fit models for the prediction of virus transmission. Mathematical frameworks that model the spread of a virus in public spaces, cities and entire countries must take into account the rate of transmission to and from each member in the population. The rate of transmission of a virus is dependent on the severity of the virus itself, population density, and mask effectiveness [10, 25–27]. The parameters relating to the effectiveness of face-masks in these transmission models vary significantly, drastically affecting the results. Eikenberry et al. included the effect of mask usage in their model based on the inward and outward efficiencies of face-masks. They cite a wide range of mask efficiencies, ranging from 20 to 80%, derived from experimental studies [15–17]. The experimental studies of course are limited in the number of subjects tested, with all experimental studies mentioned previously having less than 25 subjects and in some cases as few as 7 subjects tested, from which mask efficiencies were calculated. Eikenberry et al. illustrate the significance of proper estimation of mask effectiveness by showing that the effective transmission rate decreases linearly proportional to the mask efficiency such that masks with 20% and 80% efficiency decrease effective transmission rate by approximately 20% and 80% respectively. Such disparities in mask efficiency can lead to less than reliable models of the spread of the virus. Mittal et al. recently proposed a new transmission model, the COVID-19 airborne transmission (CAT) inequality. The model was designed for simplicity so that it can serve as a common scientific basis and also be understood by a more general audience [26]. Like Eikenberry’s and Briennen’s models, the CAT inequality accounts for the protection afforded by face coverings. The effect of face-masks in the CAT inequality is primarily based on the filtration properties of the material. All of these transmission models attempt to predict the spread of viruses, which requires a proper statistical model to account for the effectiveness of face-masks. Understanding and developing reliable models for the effectiveness of face coverings based on not only the fabric material but also the fit can lead to better transmission models. As we previously discussed, the transmission of an airborne virus depends significantly on the amount of leakage the face-mask allows.

Accurate characterization of mask effectiveness due to individual fit goes beyond reliable transmission models and more directly affects the general public. The CDC has provided design guidelines for home-made masks, but as we will show in this study the recommended mask design, or any single mask design, may not be optimally effective for the many different facial structures inside a population. That is to say, one size *does not* fit all. Instead, to ensure the effectiveness of the mask, particular sizes and designs should be recommended for several distinct population categories. In this study, we look at the leakage from home-made masks when deployed on a broad cohort of faces or population, and make suggestions on how the size and simple adjustment of face-masks can reduce the mask leakage. The study guides better utilization of face-masks to reduce transmission and can serve to reach better predictive models of the transmission of viruses such as COVID-19.

## 2 Methodology

The performance of the face-masks is highly dependent on the properties of the mask as well as the fit of the mask on a given face. Here, we employ three-dimensional morphable models of the human face to account for gender, age, and other body-habitus-associated variabilities in face morphologies and conduct mask-deployment simulations for a large “virtual cohort” of individuals. The goal is to quantify the mode of perimeter opening and to study how the mask leakage is changing with the facial features of a population. The components of the computational model are represented below.

### 2.1 Virtual Cohort of Faces

The morphable model is based on the Basel Face Model (BFM) [28], a publicly available database that includes face scans of more than 100 males and 100 females ranging from 8 to 62 years old with weight ranging from 40 to 123 kg.

Since the BFM database is pre-processed with principal component analysis (PCA), we will use the low dimensional PCA subspace to create realistic in-silico face realizations [29]. Fig 1 shows sample realizations of a face based on subspace synthesis. In addition, using identified feature vectors associated with weight, gender, age, and height, each realization can further be modified toward a particular shape. A similar morphine mesh is used for different faces, and separate regions and landmarks of the lips, ears, nose, and eyes are identified on the model. These landmarks are utilized to establish the mask position for a given face.

**Fig 1.**
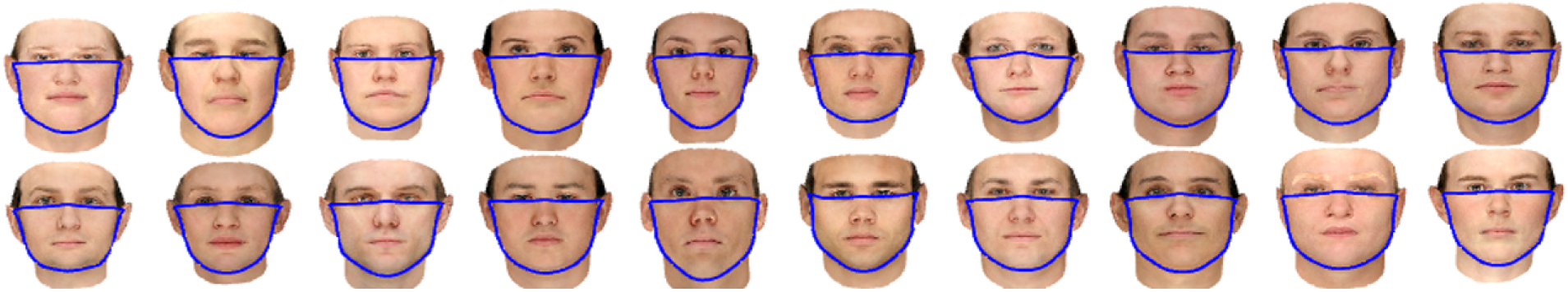
Twenty sample random realizations of the base face category from the virtual cohort.

### 2.2 Deployment Modeling

A quasi-static model is employed for the deployment of a mask on a given virtual face. In the simulation, the mask is initially placed in front of the face with the elastic bands wrapped around the ears but with zero tension. Moreover, we assumed the relaxed length of the band equals the length of the ear of each face. The resting length of the band gradually decreases from the initial length to its final value during the initial transient phases and for each stage, the intermediate quasi-static equilibrium position of the mask is calculated from the model in section 2.3. The procedure is continued until the mask rests in the final configuration on the face. The procedure is repeated for different face realizations, and for each realization, the face is systematically modified in 8 directions, namely when the face is modified toward thinner or heavier, younger or older, more feminine or more masculine, and shorter or taller features. For each case, the ensemble statistics of a particular group are calculated and cross-compared.

### 2.3 Fabric Mask Model Using Minimum Energy Concept

Because of the small flexural stiffness of the mask and geometrical constraints imposed by the band and the contact between the mask and the face, the mask could have local buckling as wrinkles and slacks on its surface [30]. To account for all of these effects, a detailed multi-scale approach is required to represent diversely scaled elements from the dominant fibers in the mask to the interaction between human facial tissue and the mask surface. Here we use the minimum energy concept as a unified principle of mechanics that works across all scales and governs the position of the mask on the face. In this method, the total elastic energy of the system is expressed as,

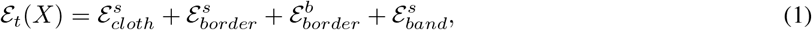

where 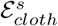 is the extensional elastic energy stored in the cloth, 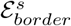 is the tension and compression energy in the border strip around the cloth mask, 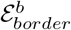 is the stored bending energy in the border strip around the cloth, and 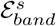 is tension energy stored in the connecting bands (refer to Fig 2 for a depiction of these effects).

**Fig 2.**
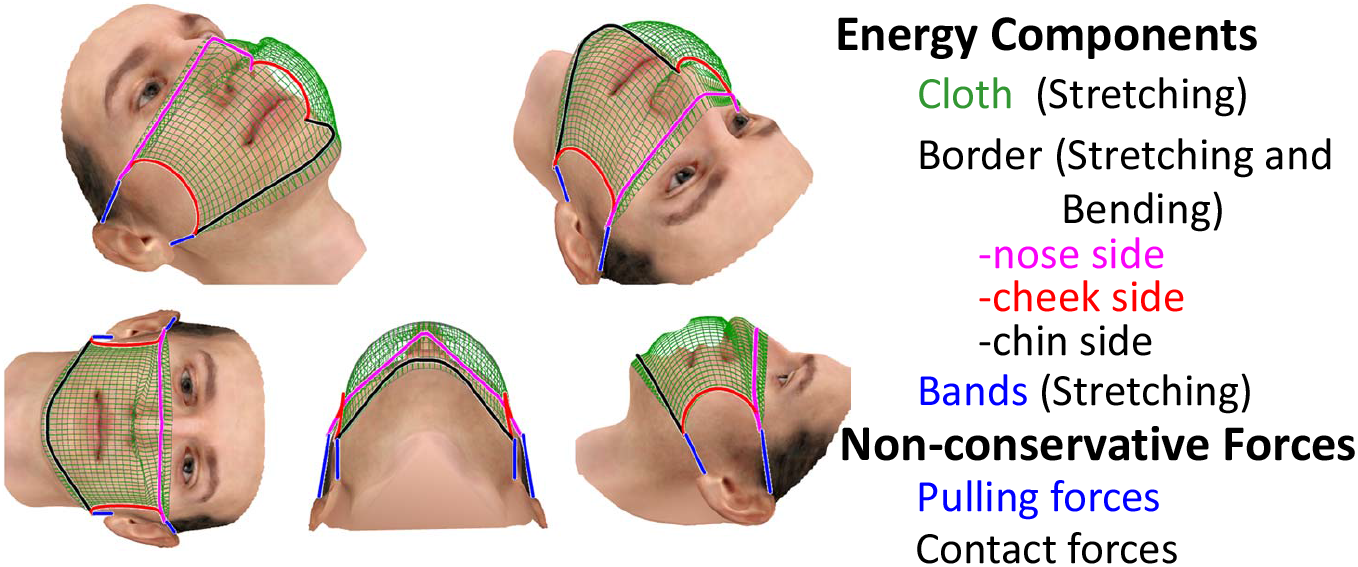
Major structural components and interaction forces of a cloth-type face-mask and their placement of the model.

The cloth is assumed to be made up of two orthonormal fiber bundles where their extensional elastic energy is an order of magnitude larger than its in-plane shear and bending energies. This assumption is justified as the regular cloth masks can be modeled as thin membranes that could easily undergo localized bucking and show negligible bending stiffness. Moreover, to account for the wrinkling effect, the energy associated with the area change of the mask is not considered. Instead, the extensional elastic energy stored in the cloth is made up of the stored energy of a group of initially orthonormal fibers according to,

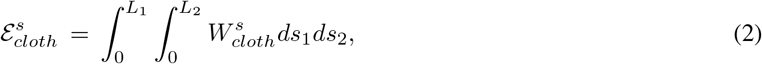

where *L*_1_ and *L*_2_ are initial unloaded edge lengths of the cloth mask. Here, 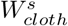 is the strain energy density defined

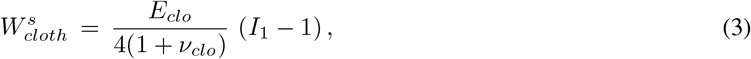

where *E*_*clo*_ is the elastic modulus and *ν*_*clo*_ is the Poisson ratio of the cloth respectively, and *I*_1_ = 2(*D*_11_ + *D*_22_) is the first invariant of Green strain tensor defined as,

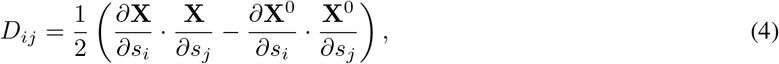

with **X** and **X**^0^ are the current and initial position of the cloth, respectively.

Similarly, 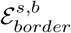 is defined as

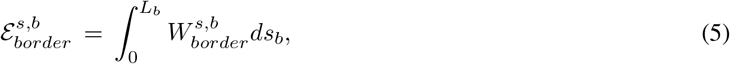

Where 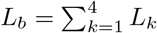, is the summation of the length of all four edges of the cloth mask. Here, if **X**(*s*) and **X**^0^(*s*) are the coordinates of the border in the reference and deformed configurations respectively, we can define the attached coordinate system to the border in its deformed configuration using its tangent vector 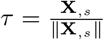, binormal vector 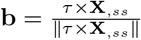 and normal vector 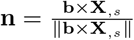. A similar definition is also used for the reference position of the border. The extensional strain energy density functions can then be expressed as

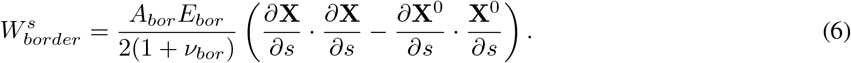

The bending strain energy density function is approximated as

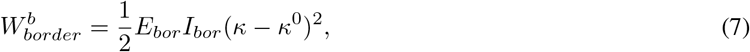

where *κ* and *κ*^0^ are the curvature in the direction **n** and **n**^0^, respectively. Here, the initial curvature of the border is chosen to be zero. The curvature can be related between local sets of three consecutive discrete nodes **X**_*i*−1_, **X**_*i*_, **X**_*i*+1_ along the rod according to [31],

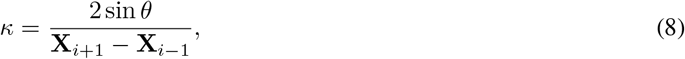

where *θ* is the angle between two consecutive segments of the line and is defined as,

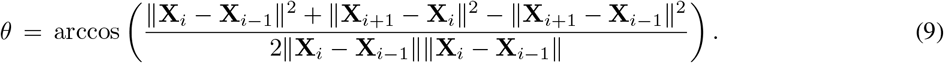

The energy contribution from the stretching band is also defined similarly to 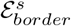, with the extensional stiffness of 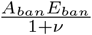.

In addition to internal energy action, the non-penetration contact force between the mask and the facial tissue is represented with non-conservative forces, **f**_*t*_ = **f**_contact_, acting on the mask surface. Here, we assume soft contact between the face and the mask, in which *f*_contact_ is defined as,

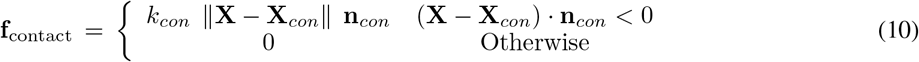

where **X**_*con*_ is the closest point on the face to point **X** on the mask, and **n**_*con*_ is the outward normal to this point. The contact stiffness between the skin and the mask is represented by *k*_*con*_.

By relating the internal forces of the mask to the derivatives of the energy density function, nonlinear sets of equations are obtained for the node placements *X*_*k*_ = (*x, y, z*)*k* in the discrete model of the mask. The resulting equations are solved to find the equilibrium position at a given deployment stage. Moreover, since the equilibrium shape is slowly modified from the previous deployment stage, **X**^*n*−1^, a linearized equilibrium equation is derived for *δ***X**=**X**^*n*^ **− X**^*n*−1^, and is employed as a preconditioner to accelerate the convergence of the solution. This is done by defining spatial virtual work in terms of virtual velocity, *δ***v** (**X**), and a discretized solution of **X**, *ϕ*(**X**), obtained from the discrete models of the mask, border and band. The equilibrium equation is solved iteratively to find the new position **X**^*n*^ from **X**^*n*−1^ using the projective dynamics methodology [32].

Without loss of generality, here we assume that *E*_*clo*_ = *E*_*bor*_ = 10 Mpa and the band is 4 times stronger with *E*_*ban*_ = 40 MPa. All the Poisson ratios are fixed at 0.3 [33]. The thickness of the cloth mask is chosen to be 0.5mm, and the band is made up of 0.5in folded cloth fabric following the recommendation of the CDC for the short edges [34]. The stretching band is assumed to be circular with a diameter of 1mm. These parameters are chosen to be close to typical cotton fabric and elastic band. In addition, it is assumed that the contact stiffness between the mask and different part of the face is similar at every location on the face with the contact stiffness *k*_*con*_ = 1 Mpa, in the range of skin (0.6 MPa) and thick muscles (∼ 0.8 MPa) [35, 36]. Also, the cloth mask is discretized with Δ*s* = 2*mm*, and grid refinement studies have been performed to ensure that the simulation results are independent of the grid sizes.

## 3 Results

### 3.1 Effect of Mask Size and Side Tuck-in Ratio

Here, the leakage area for a rectangular cloth face-mask for three different tuck-in conditions of the side edges is explored. The tuck-in of the mask is mathematically modeled as the gradual reduction of the unstretched length of side edges from the initial length (*L*_0_) to *L* = *α*_*T*_*L*_0_, where *α*_*T*_ is named as the tuck-in ratio (Fig. 3). The mask shape and size are chosen based on the guideline by CDC on how to sew cloth face coverings [34]. From the guideline, the baseline case is selected to be a mask with *L*_0_ = 5.5*in* and *W* = 9*in* (will be referred to as medium mask), with *α*_*T*_ = 0.5. Two other sizes of *W* = 8*in* (hereafter will be referred to as small mask) and *W* = 10*in*. (will be referred to as large mask) with the same aspect ratio are also tested to account for the variation in the mask size.

**Fig 3.**
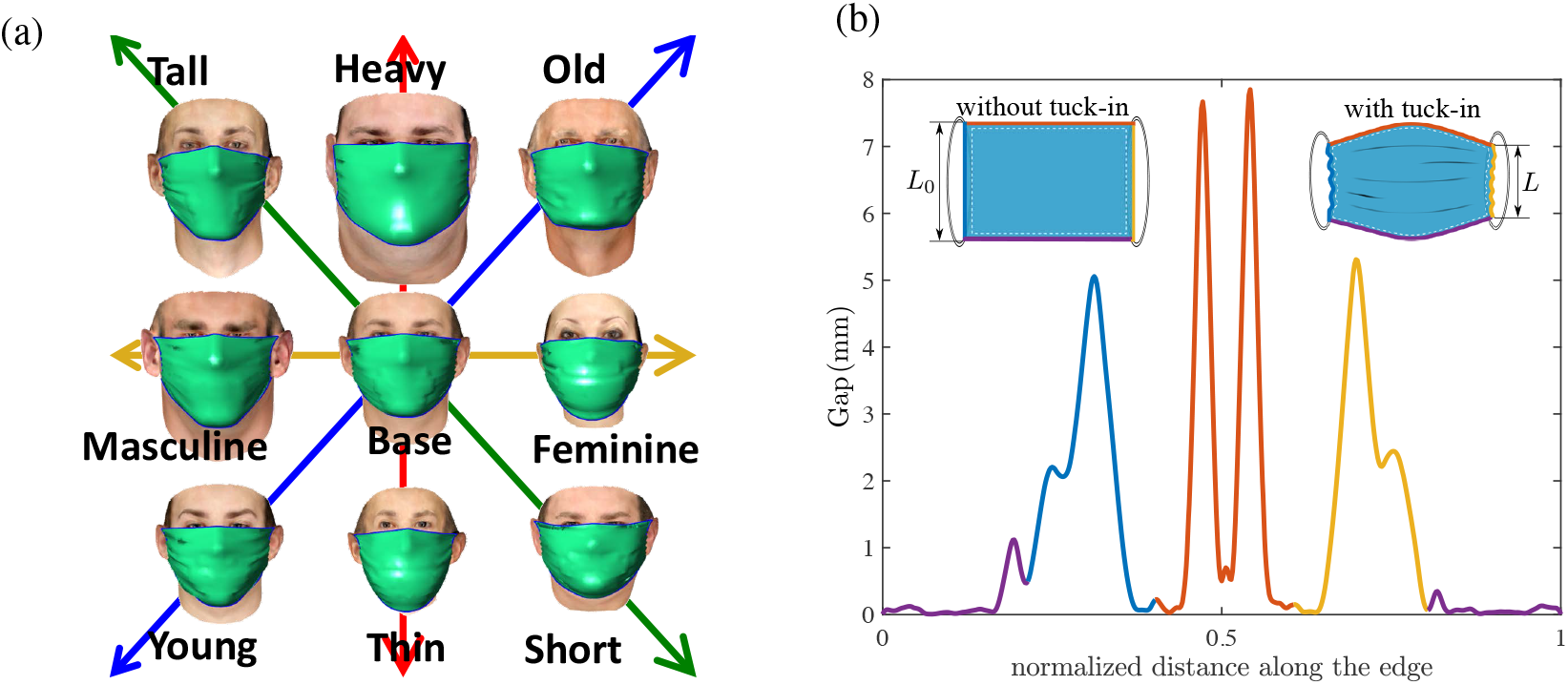
a) Sample simulation results of deploying a face-mask on a base realization face and its modification along each feature axes. b) A sample profile of leakage opening around the perimeter of the mask in the clockwise direction starting from the middle of chin side. The mask edges are named as the nose side (Area 1), cheek side (Area 2) and chin side (Area 3) and are marked with their corresponding colors in the inset figures. The inset figures also shows the definition of tuck-in ratio *α*_*T*_ = *L*/*L*_0_ in the current study.

To ensure the ensemble statistics are sufficient to reach a confident inference from the simulations, 150 random subjects are selected from the virtual cohort of faces and for each subject, 8 modified configurations are generated to systematically explore how the facial features affect the leakage area around the perimeter of a mask. The modification to each random face is done along one feature direction at a time. The feature vectors in this study are restricted to weight (thin to heavy), age (young to old), gender (feminine to masculine), and height (short to tall) as shown in Fig 3a. The selection is based on the availability of the prior database. Other important features of faces such as race are not considered and will be explored in future work, as explained in the conclusion section.

Fig 4a shows the statistical mean value of the cumulative leakage area around the perimeter of small, medium, and large masks with the tuck-in ratio of *α*_*T*_ = 0.7, 0.5, 0.3. The bars are for the CDC recommended mask (medium size) while the blue and red dots represent large and small masks, respectively. Each category of faces are plotted with a different color and the tuck-in ratios are shown with bars with solid (*α*_*T*_ = 0.7), dashed (*α*_*T*_ = 0.5) and dotted (*α*_*T*_ = 0.3) borders. The error-bar for each data shows the standard deviation of the computed parameters for the 150 random face realizations. It is found that the smaller mask size, relative to the CDC recommended size, has minimal effect on the total leakage area for the base cases, especially for higher tuck-in ratios. However, there are substantial changes in the total leakage area for thinner, younger and more feminine faces with a decrease in mask size regardless of the tuck-in ratio. This suggests that recommending one mask size for the entire population and recommending a tuck-in adjustment can only be effective for certain categories of faces and will not substantially improve the protection of a broad category of people.

**Fig 4.**
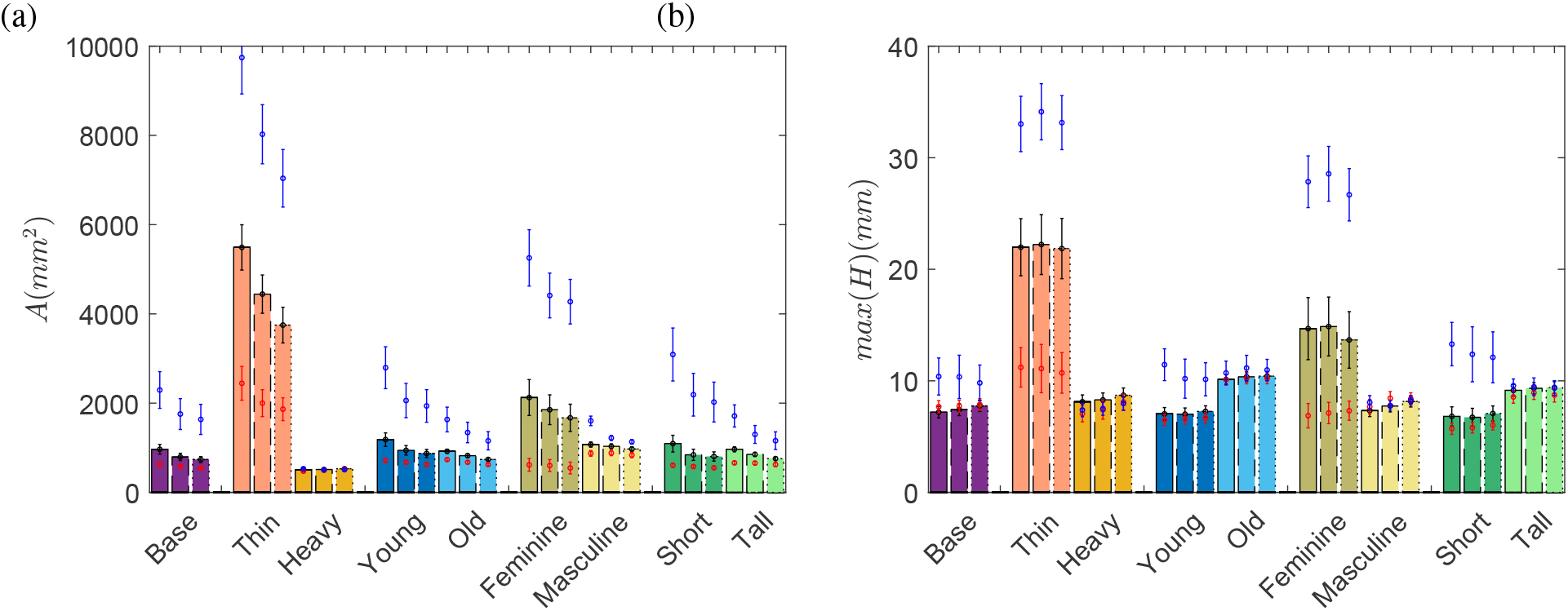
a) Average perimeter leakage opening area, b) the maximum gap distance. Bars are for medium (CDC recommended) mask. Red and blue dots represent the small and large masks, respectively. The tuck-in ratio of 0.7 is shown with solid bars, 0.5 with dashed bars, and 0.3 with dotted bars. The standard deviation of each data point is displayed with a line.

Except for the heavy category, the total leakage area continuously reduces with more tuck-in on sides. The reduction rate is not present when the mask is small. Based on these results, we suggest that different mask designs should be recommended for different categories of people, especially based on weight, age, and gender. It is also found that feminine and thin faces can substantially benefit from the smaller masks. Also, results suggest that the weight can serve as a critical indicator for the recommendation of optimal mask shape.

The leakage around the edges of the mask is also dependent on the hydraulic perimeter of the opening (both area and shape of opening) [37, 38]. To explore this effect, in Fig 4b we show the maximum opening (maximum distance between the mask edge and face). Surprisingly, except for thin, feminine, and short faces, the reduction of the mask size from the CDC recommended dimensions does not substantially reduce the maximum gap opening (Fig 4b).

However, the maximum gap can be affected by the tuck-in ratio and, more importantly, by the facial features. As an example, smaller tuck-in ratios result in larger gap distance in older faces. The largest difference in the maximum opening is found between the thin and heavy categories, followed by feminine and masculine faces. The main observation is that proper mask sizing is the most effective way to reduce the maximum opening and a smaller tuck-in ratio could only be beneficial in certain categories of faces.

The comparisons between mask sizes and tuck-in ratios for the median cases in each category are shown in Fig 5. It can be seen that the placement of the mask on the face is greatly modified when the mask becomes smaller than a threshold. In particular, the lower edge of the mask shifts from below the chin to the top of the chin for heavy and tall faces with a small mask. Consequently, the bottom support of the mask can slip easily in tangential directions and could easily lead to changes in the mask placement during routine daily activities such as talking and breathing. The thin and feminine cases exhibit a rapid increase in the opening gap, primarily in the chin area, with an increase in the mask size. The results suggest that addition of a tuck-in mechanism to the lower edge of the medium size mask is a simple modification that would make them more effective for feminine and thin faces. The adaptation of a smaller tuck-in ratio on the side-edges, on the other hand, leads to local increases in the maximum opening gap without changing the opening area noticeably.

**Fig 5.**
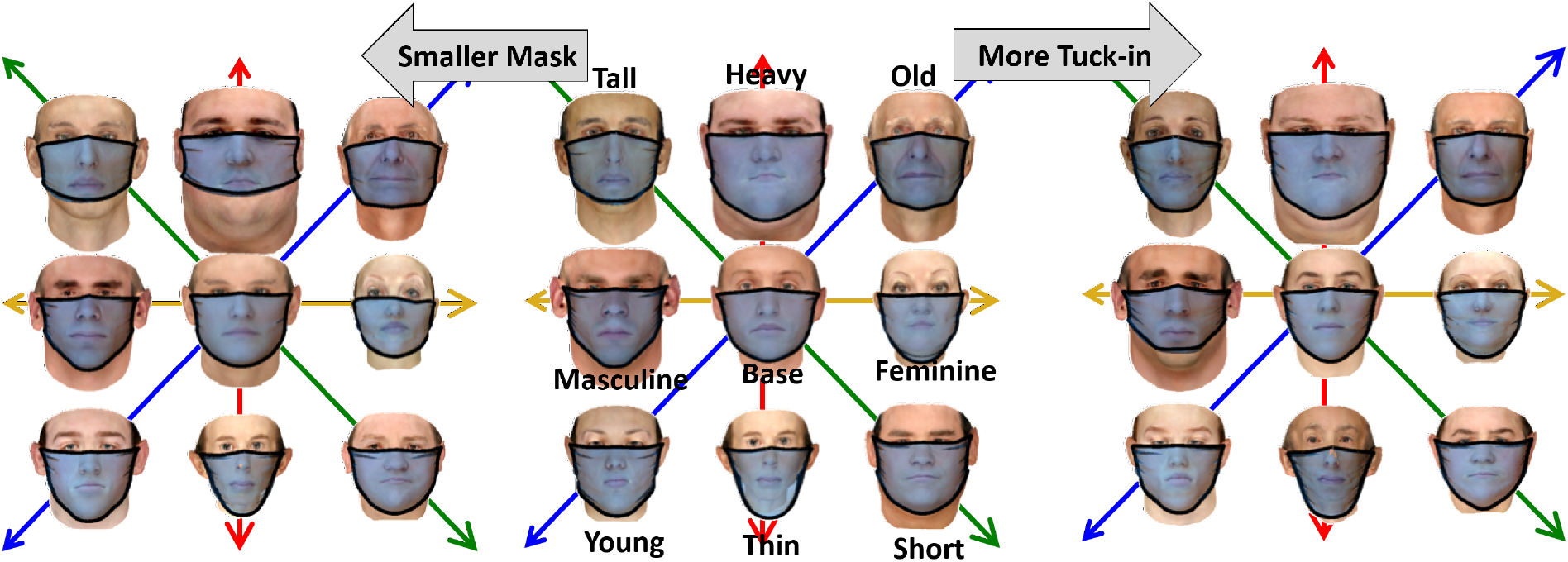
The middle plot are the median cases for the medium mask (CDC recommended size) and tuck-in ratio of 0.5. Right and left plots show the modification for the small mask and tuck-in ratio of 0.3, respectively.

The leakage area and maximum gap around the mask can be divided into three distinct segments: the top edge near the nose, the side edges near the cheeks, and lower edge near the chin. Edges’ names and locations are shown in Fig 3c. The maximum gap opening and the contribution of each segment of the mask to the total leakage area are compared in Fig 6. It is observed that the leakage from the nose area is independent of the tuck-in ratio and mask size, except for thin, feminine, and to some extend, short faces and the large mask. When the mask is medium or small, the tuck-in ratio can be used to further reduce the opening near the nose (side 1) in thin and young faces without increasing the maximum gap distance. For the other cases, lower tuck-in ratios are mostly accompanied by an increase in the maximum gap opening on top of the mask. The increase is primarily due to changes in the placement of the mask on the nose and is more pronounced in masculine and heavy faces. The maximum gap opening from side 1 in older faces is insensitive to the mask size or tuck-in ratio, suggesting that a clip or other mechanical attachment is necessary for this category to reduce the gap opening.

**Fig 6.**
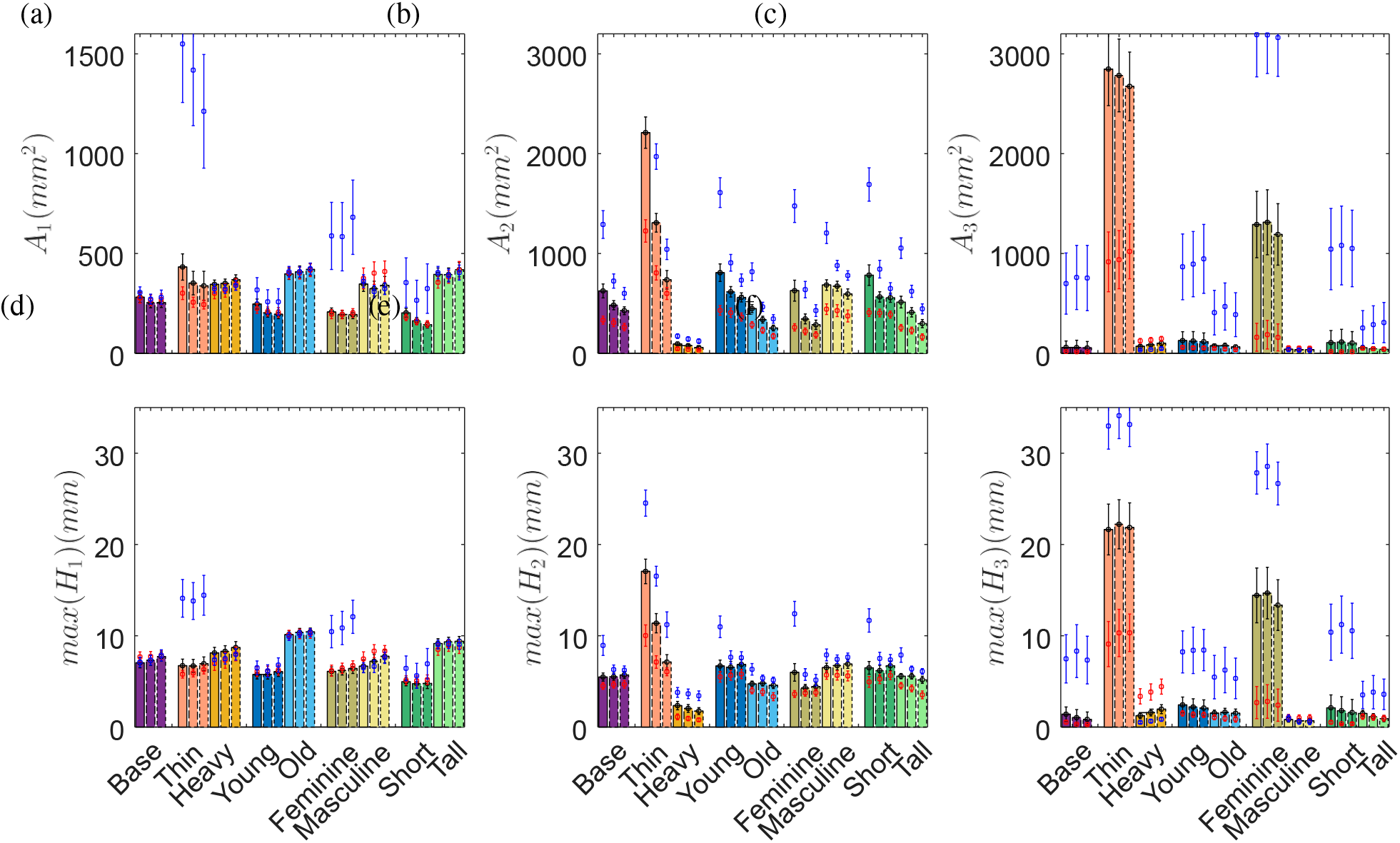
Average perimeter opening area (a-c), and maximum opening gap (d-f) in three edges of the mask as marked in Fig. 4c. The description of the data is similar to Fig. 4.

A different trend is observed for the cheek edges (side 2) where there is a substantial reduction of opening with a decrease in mask size and tuck-in ratio. Nevertheless, except for the thin faces, the maximum observed gap is similar for larger mask sizes and all tuck-in ratios. The leakage area from this side shows the greatest reduction with smaller tuck-in ratios relative to the other two regions of the mask. The reduced area and unchanged maximum opening indicate that with smaller tuck-in ratios, the side opening gap becomes more concentrated while the top amplitude of the gap profile does not change substantially.

The maximum opening and the leakage area near the chin (side 3) are major components of the total leakage for thin and feminine faces. The tuck-in ratio has an insignificant role in this part of the mask, while the mask size is the primary driving factor. Thin and feminine faces show a more than 50% reduction in leakage area with the smaller than the CDC recommended mask size. An exception is observed for heavy faces, where a small mask induces larger maximum openings in the lower edge due to the slip of the face-mask on the face and placement of the lower edge on top of the chin area.

The previous results show that both leakage area and maximum gap opening of the edges should be considered to reach a discriminatory factor that can identify different modes of leakage around the mask. Toward this, we found that 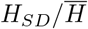 can serve as the parameter for unsupervised classification of the results, where *H*_*SD*_ is the standard deviation of the opening gap and 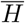 is the average opening distance along the edges. Fig 7 shows the scatter plot for mask size and tuck-in ratio effects. It is found that the data can be grouped into 5 clusters to best separate the effect of facial features. The number of clusters (*K*) is chosen such that with a further increase of *K*, the reconstruction error is not significantly reduced. The reconstruction error is defined as 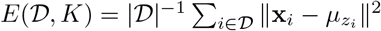, where 𝒟 is the data set and *μ* is the center of the cluster 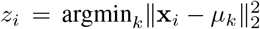. These clusters are marked with different colors in each sub-figures, while different symbols are used to distinguish between different feature categories. Besides, the inset figures present the percentage of face categories in each cluster.

**Fig 7.**
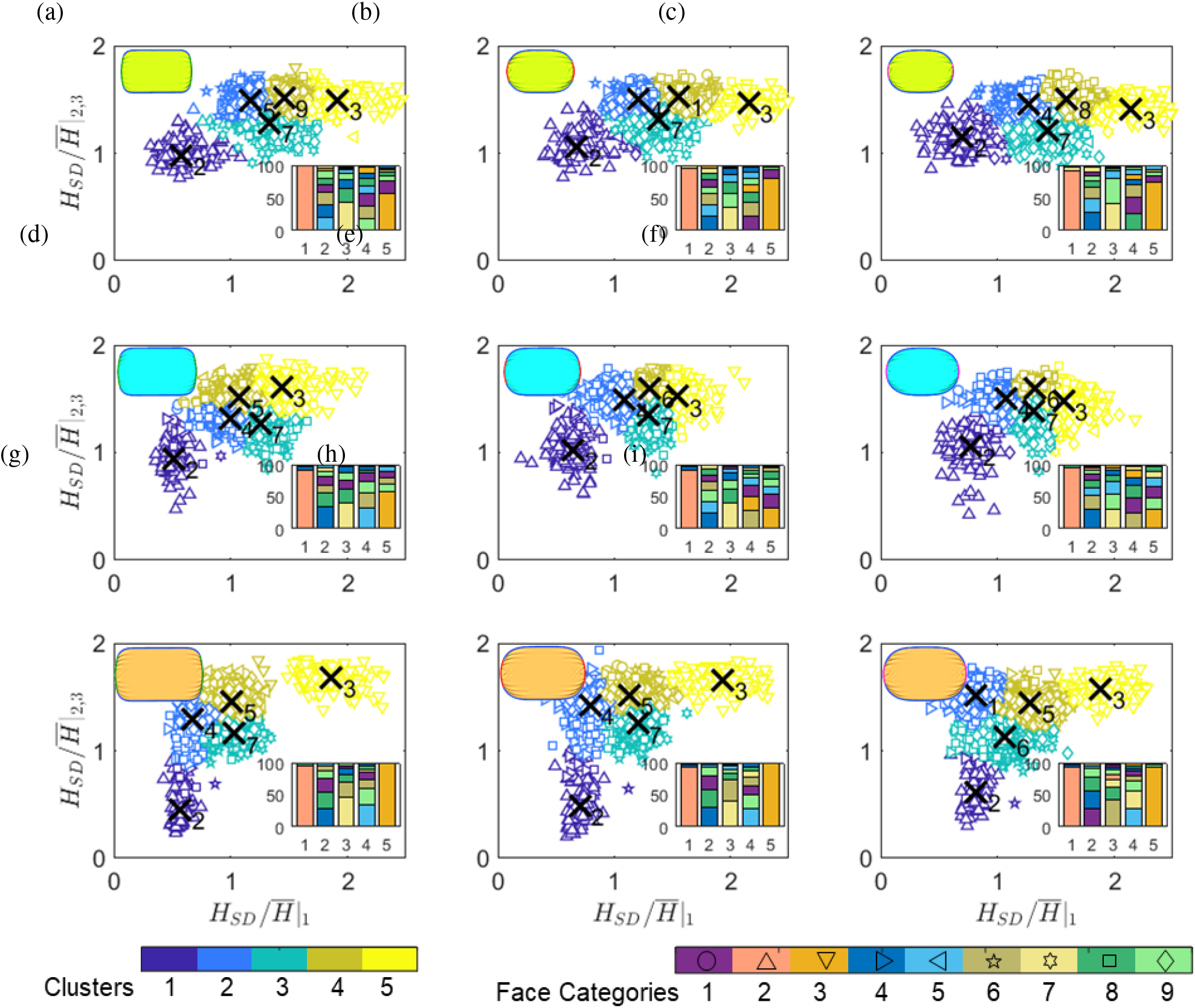
Classification of the leak for different mask size (rows) and tuck-in ratio (columns) based on the shape parameter of 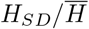. Clusters are displayed with different colors and different face categories are marked with symbols given in the legend. The inset figures represent the percentage of constituent faces in each category.

The thin face category (number 2 in the legend) is consistently the primary contributor of cluster 1 (dark blue cluster). Similar observations can be made for heavier faces and cluster 5 (yellow cluster). The center of cluster 1 shifts to higher 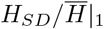 and 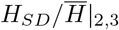 with decreasing tuck-in ratio, consistent with previous observations that a smaller tuck-in ratio leads to more non-uniform gap distributions. An increase in mask size shifts the cluster centroid to lower 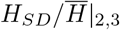, indicating that the mask size will primarily affect the gap opening on lower and side edges. Cluster 5 only shows variations along the 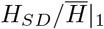 axes. Also, we see that the masculine face category is the most prevalent member of cluster 3, with a minor centroid shift among the cases. The other two clusters, clusters 2 and 4, are mixed sets of faces suggesting that other facial features are needed to classify this region of the features sub-space. It is found that the tuck-in ratio can only induce a minor shift to these clusters, but the mask size can substantially modify the mode of opening along the edges.

In Fig 8, we plot the cases of the dominant feature category nearest the cluster centroids of Fig 7 for medium (a), small (b), and large (c) mask sizes. Each figure also include the results for the tuck-in ratios of 0.7 (top row), 0.5 (middle row) and 0.3 (bottom row). As mentioned previously, clusters 1, 3, and 5 are predominantly comprised of the thinner, more masculine, and heavier faces, respectively. The young and short faces are the most representative feature categories of cluster 2, while the old and feminine faces form the majority of cluster 4 for medium and large masks. Nonetheless, it is found that none of the feature categories is the dominant constituent of clusters 2 and 4 with more than 1/3 of the members.

**Fig 8.**
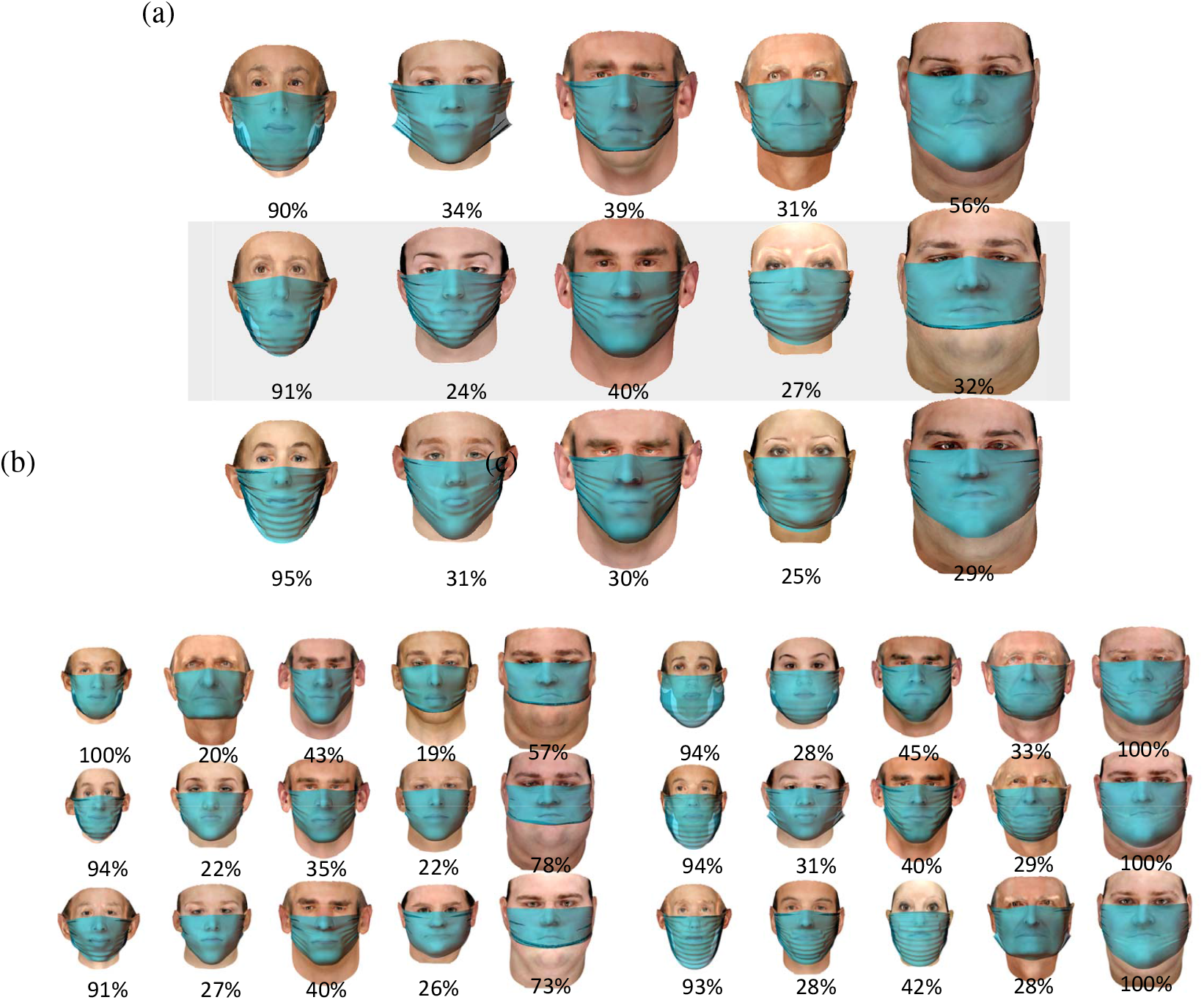
The nearest cases to the center of the clusters and the percentage of them in each cluster for (a) medium(CDC recommended) mask, (b) small mask, and (c) large mask for three tuck-in ratios of 0.7,0.5 and 0.3 from the top row to bottom.

The results from unsupervised clustering based on the face-associated variabilities suggest that for certain groups such as heavy and thin faces, it is possible to find the most effective face covering with minimal gap opening, especially with the use of proper mask size. However, for the other cases, one needs to consider other factors such as shape and geometrical parameters of the face to better identify the most optimal cloth mask covering size and tuck-in ratio.

### 3.2 Role of Facial Features

In this section, we explore how the changes in the categorical facial features (weight, age, and gender) affect the findings presented. The leakage area and maximum gap distance are shown in Fig 9. The horizontal axis is hereafter named as weight/age/gender index, with a zero value corresponding to the base case. The associated facial feature for each value is depicted in the legends, and the cases used in the previous sections are marked with. The left column is for different mask sizes and the tuck-in ratio of 0.5 and the right column is for the medium (CDC recommended) mask size with tuck-in ratios of 0.3, 0.5, and 0.7. The dash lines are showing the standard deviation of data. Finally, in Fig 8d we plot the median cases for the marked dots in the sub-figures (i-1) for the medium mask and the tuck-in ratio of 0.5.

**Fig 9.**
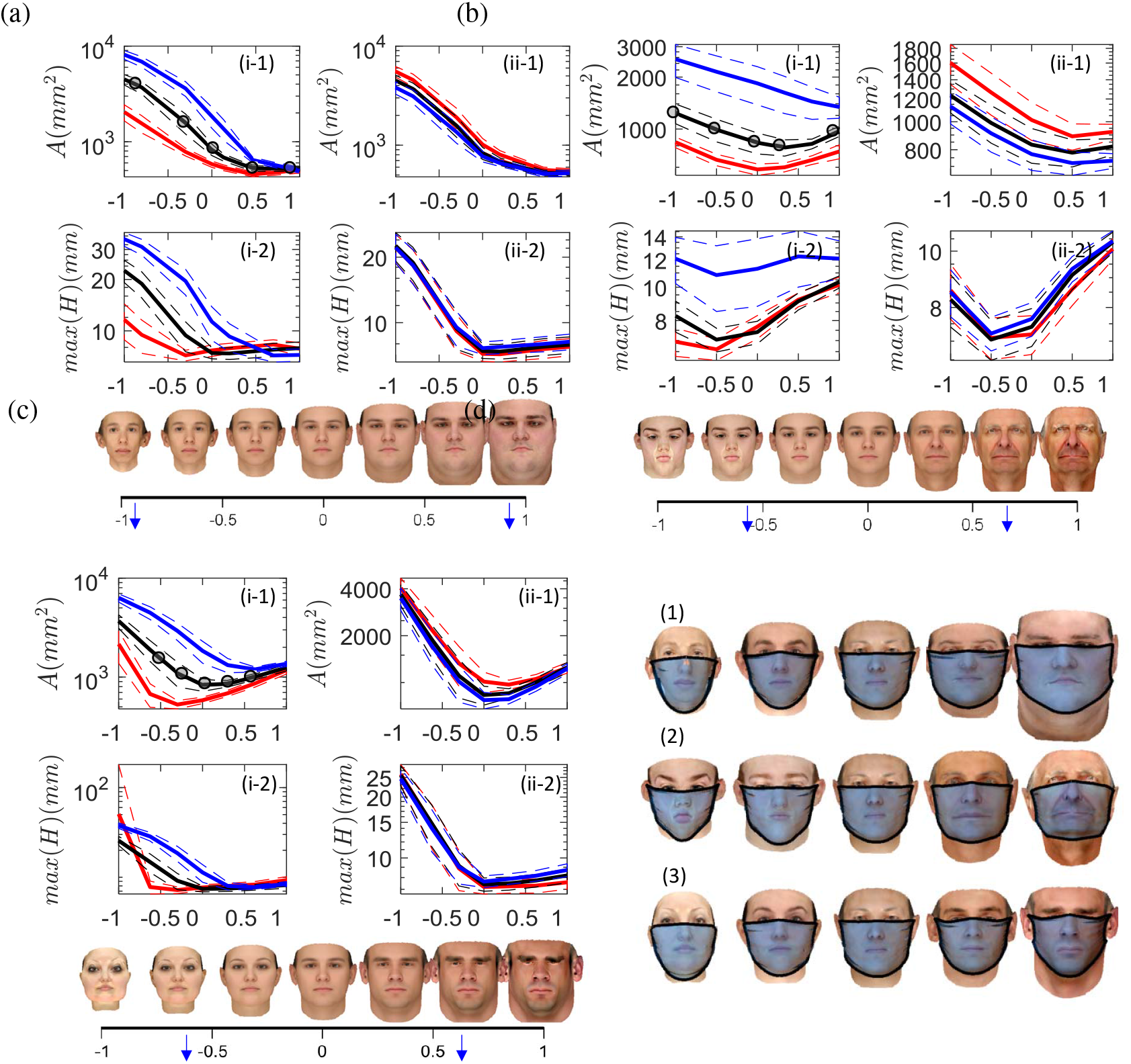
The changes in the leakage area (part 1), maximum gap opening (part 2) for different (a) weight, (b) age and (c) gender indices. Panel (i) is for 3 mask sizes (red: small, black:medium, and blue: large) with tuck-in ratio of 0.5, and panel (ii) is for 3 tuck-in ratio (red:0.3, black:0.5 and blue:0.7) and the medium mask size. The x axis is the normalized feature mode shown for the average face in the subset figures. The median cases for the medium mask size and tuck-in ratio of are 0.5 marked in the subfigures i-1.

The increase in the weight feature results in an exponential decay in the leakage area until a threshold at which the leakage area reaches its asymptotic value. This threshold is very similar between different mask sizes and happens at a weight index of 0.5 (Fig 8a-i). The weight index with the minimum gap is highly dependent on the mask size, observed at −0.3, 0, and 0.6 for the small, medium, and large masks respectively. On the other hand, the higher tuck-in ratio marginally reduces the opening area for all weight indices and has almost no effect on the minimum opening gap (Fig 8a-ii panel). As shown in Fig 8d-1, the gap opening along the bottom edge of the mask changes most significantly and its tightness on the chin is correlated with the transition point observed above.

The change in the age feature of the face has a different outcome on the leakage area and maximum gap opening. Both the tuck-in ratio and mask size almost equally modify the leakage area and maximum opening with the age index (Fig 8b). The minimum leakage area with respect to the age index shifts to older faces with increasing mask size but is not affected by the tuck-in ratio. The large mask shows almost similar maximum opening across all ages, while the other sizes show initial decay and subsequent rise with age index. The smallest maximum opening occurs at a lower age index than the does the smallest leakage area. The tuck-in ratio further changes the maximum opening, especially at the low and high extremes of the age index. The median realizations for highlighted cases in panel (i-1) in Fig 8d-2 indicate that there is a shift in the location of the upper edge of the mask on the nose with the age index. A simple mask design is found to be incapable of reducing the top edge opening near the nose as the geometric dissimilarities between the mask and the face always lead to an asymptotic opening.

The gender index, while showing a similar trend in total leakage area to the age index, has a distinct maximum opening profile, which is more similar to that of the weight index (Fig 8c). The minimum leakage area is shifted to more feminine faces with smaller mask sizes, but the maximum opening becomes larger with increase of feminine gender index. In fact, the smallest mask exhibits the largest gap opening than any other case when the gender index is −1 (most feminine). For more masculine faces, the response of all mask sizes is similar. Finally, the tuck-in ratio has negligible effects across the gender index. From the tested cases, it is found that all gender faces have similar gap distributions in cheek and chin areas, but they are different in the opening at the nose area compared to other indices like weight.

## 4 Discussion

The findings from the previous sections show that cases with smaller facial dimensions such as thinner, younger, and more feminine faces, tend to suffer from more leakage due to improper fit. There is, in fact, a threshold in mask size at which these faces show a significant increase in leakage area, especially from the bottom edge of the mask. Based on the depiction of the median cases (Fig 9d), it is observed that this increase is due to an oversized mask hanging off the face near the chin. Of the three mask sizes studied, thin and feminine faces were best served by the small mask, which compared to the CDC recommended size reduced the leakage area by over 50%. We also found that heavy and masculine faces could be at a disadvantage with smaller masks as small masks could slip above their chin and increase the risk of new perimeter leaks during routine daily activities like breathing and talking. The other simple modification to masks, besides the size, is the tuck-in of the side edges of the mask. In general, larger tuck-in (smaller tuck-in ratio) leads to reduced leakage areas. This change is mostly noticed for the smaller faces i.e. thin and feminine faces. Compared to the mask size parameter, though, tuck-in does not greatly affect the leakage area.

A proper fitting mask should reduce the leakage area as much as possible. However, as mentioned in section 3.1, the shape of the opening is also crucial in determining the effectiveness of a mask, especially for outward protection. We describe this with the maximum gap opening *max*(*H*). Given the same leakage area, the mask that produces larger *max*(*H*) has more localized openings as opposed to the mask with smaller *max*(*H*) would have a more uniform slit-like opening through the length of the mask edge. During a respiratory event, the more localized openings would create higher exit velocity jets, which would in turn, spread the aerosols further. From this, we cannot definitively conclude that masks that reduce leakage areas are better than the CDC recommended mask. Instead, we must also ensure that the reduction in leakage area is not accompanied by an increase in *max*(*H*). Except for the heavy and masculine faces, reduction of mask size led to a decrease in *max*(*H*). We already discussed that mask size reduction does not provide any added benefits to heavy and masculine faces; now, we see that smaller masks are detrimental to the overall mask effectiveness in these categories of faces. Previously we mentioned that more tuck-in has a minor effect of the leakage area in most categories. However, except for thin, feminine, and short faces, increased tuck-in leads to larger gap openings.

The weight factor shows the most effects of the modifications of the mask design. A study of the direct effect of the weight-dependent facial features shows that after a weight-index of 0.5, mask size and tuck-in have no effect on the leakage area. We already established that thinner faces (weight-index < 0) benefit from the small mask and tuck-in. Heavier faces were observed to have more uniform openings (smaller maximum gap) with the larger masks but this was greatly dependent on the weight index. The same analysis was carried out on the age and gender feature categories. Both of these saw, by and large, a decrease in leakage with smaller masks across the feature’s index. Masculine faces, similar to heavier faces, show negligible effects of both mask size and tuck-in ratio. The feminine faces did show some reduced leakage area when the mask is small but interestingly, the small mask produced the largest maximum gap opening in the most feminine faces (index of −1). For both the age and gender indices, there appears to be an optimal index where the leakage area and maximum opening attain their minimum values. This would lead one to believe that there are parameters other than the feature categories explored here, on which the mask fit depends. We have alluded to one such parameter already through this study, the face size.

## 5 Conclusion

The effect of mask fit for a large cohort of individuals with varying facial features was studied using three-dimensional, morphable, headform models onto which an elastic mask was deployed via a quasi-static simulation. The categorical study of facial features (weight, age, gender, height) prove that the CDC recommended mask size is perhaps not the most effective mask size for the entire population. Thin, young, feminine, and short faces were observed to benefit from a smaller mask size. Heavier and masculine faces on the other hand, would benefit, albeit very little, from a larger mask. More importantly, tuck-in of the side-edges alone cannot reduce the leakage area considerably and, in turn, can cause larger gap openings. The effect of the tuck-in ratio on the sides of the masks was observed to be beneficial only on thin, young, feminine, and short faces. However, for the remaining categories, tuck-in was generally associated with minimal change of the leakage area and a substantial increase in the maximum gap opening.

From this study, it is clear that subjects with smaller faces, such as thin or feminine faces, should use smaller masks with side-edge tuck-in. Also, a lower edge tuck-in modification could reduce the leakage near the chin, which is the principal contributor to the total leakage area in these subjects. We would further recommend, based on the observations made here, that except those with heavy and masculine faces use smaller than the CDC recommended mask size. Side edge tuck-in for inward protection is marginally effective across all categories. However, based on this study alone, no conclusion can be made on the effect of tuck-in for outward protection due to the shape change of the openings. Analysis on the flow of respiratory events needs to be carried out to arrive at a definitive conclusion on this. Besides a general increase of the maximum gap opening at the nose with tuck-in, the perimeter leakage from the top edge of the mask was unaffected. A wire strip at the nose such as that implemented in other masks (e.g. surgical masks and N95 respirators) are recommended to reduce the top edge leakage. The aspects discussed provide guidance for recommending simple mask designs based on easily accessible body-habitus features to improve public safety during COVID-19 and future similar pandemics.

As a future direction, more facial features should be included in the population-based study. In the current study, an important factor of the race is not considered mainly due to the availability of the prior data used for creating the virtual cohort of faces. Also the comfort factor is another detrimental factor for wearing a face mask furing daily activities. The study should be extended to include these factor as well to form a holistic predictive measure that can be employed in airborne disease transmission models for accurately diverse populations. Moreover, the leakage area by itself might not be a complete descriptor of the inward and outward risk of the mask. The flow speeds during inspiration and expiration phases are very different. It is anticipated that lower pressure in the region interior to the face-mask during inhalation could induce inward deformation to the mask and therefore reduce the perimeter leaks. On the other hand, the higher pressure during the expiration process might create larger leakage openings and depend on the instantaneous shape of the mask, induce stronger or weaker leakage jets on sides. These factors will be explored in future studies.

## Data Availability

All the required data is provided in the paper

## Acknowledgment

We would like to acknowledge Drs. Jung-Hee Seo and Kenny Breuer for inspiring discussions and feedbacks on this study. We also acknowledge the Florida State University Research Computing Center and the National Science Foundation XSEDE programs for the computational resources that made this study possible.

